# Findings and prognostic value of lung ultrasound in COVID-19 pneumonia

**DOI:** 10.1101/2020.06.29.20142646

**Authors:** Jorge Castelao, Desirée Graziani, Collaborative Working Group COVID Pulmonology Guadalajara, Joan B. Soriano, José Luis Izquierdo

## Abstract

**Objectives:** The aim is to systematically describe the findings of lung ultrasound in patients with COVID-19 pneumonia and to analyze its prognostic value.

**Methods:** Lung ultrasound was performed to 63 patients with COVID-19 pneumonia admitted to a University Hospital. Lung involvement was evaluated using a 4-point scale with a 12-area pulmonary division (lung score -LS-). Ultrasound findings, along with clinical characteristics, were recorded.

**Results:** All patients showed ultrasound involvement in at least 1 area (mean 8 ± 3.5). Total LS was 15.3 ± 8.1, without differences between left and right lung. Most affected regions were the lower one (95.2%) and the posterior one (73.8%). Total LS showed a strong correlation (r = -0.765) with PaO_2_/FiO_2_; by lung regions, those with a higher correlation were the LS of the anterior one (r = -0.823) and the LS of the upper one (r = -0.731). 22.2% of patients required non-invasive respiratory support (NIRS). Multivariate analysis shows that anterior region LS, adjusted for age and sex, is significant (odds ratio 2.159, 95% confidence interval 1.309 to 3.561) for the risk of requiring NIRS. Anterior region LS ≥ 4 and total LS ≥19 have similar characteristics to predict the need for NIRS.

**Conclusions:** Ultrasound involvement in COVID-19 pneumonia is bilateral and heterogeneous. Most affected regions are the posterior and the lower ones. The anterior region has prognostic value, because its involvement strongly correlates with the risk of requiring NIRS, and an anterior region LS ≥ 4 has high sensitivity and specificity for predicting the need for NIRS.

## INTRODUCTION

In December 2019, a new coronavirus was identified in hospitalized patients in Wuhan (China)^1^, initially named 2019-nCov. On February 11, 2020, the World Health Organization (WHO) formally named COVID-19 the disease caused by this new virus, whose name changed, by decision of the Coronavirus Study Group of the International Committee for the Taxonomy of Viruses, to SARS-Cov-2. The number of COVID-19 cases grew exponentially and on March 11, 2020, the WHO officially declared the pandemic^2^. As of May 20, 2020, the WHO recognizes 4,731,458 cases worldwide, and 316,169 deaths^3^. On the same date, in Spain, the Ministry of Health acknowledges the (official) existence of 232,037 cases and 27,778 deaths^4^.

COVID-19 symptoms are mostly nonspecific and common to any respiratory tract infection. Approximately 14% of patients develop severe disease and 5% present a critical situation^5^.

Imaging tests are necessary to evaluate patients with suspected pneumonia. Pulmonary ultrasound is a portable, fast, radiation-free, non-invasive method that has been used for the initial assessment, diagnosis, prognostic stratification, and monitoring of pneumonia in the pre-COVID-19 era^6,7^ and during the COVID-19^8-10^.

First to report lung ultrasound results in patients with COVID-19 were Huang et al^11^, with a non-systematic description of findings in 20 patients. Since then, numerous works have been published, but most are case descriptions, small series, or recommendations and reviews^8-19^. All of them are based on the search for classic ultrasound signs of interstitial involvement (B-pattern) and consolidation (C-pattern), originally described by Lichtenstein^20,21^.

Numerous ultrasound findings have been reported in patients with COVID-19, but a systematic description of the ultrasound presentation of COVID-19 pneumonia has not been published to date.

The aim of this work is to describe in a systematic way the pulmonary ultrasound findings in patients with COVID-19 pneumonia, and to analyze if echographic characteristics correlate with pneumonia severity and/or with clinical course.

## PATIENTS AND METHODS

Prospective study carried out in patients admitted with the diagnosis of COVID-19 pneumonia in a University Hospital that serves a population of 245,000 people. The project has been approved by the Clinical Research Ethics Committee of the University Hospital of Guadalajara (Spain), and informed consent has been obtained from all patients.

From March 19, 2020 to April 19, 2020, thoracic ultrasound was performed on 63 patients admitted to the Pneumology Service (no patients were excluded) with a diagnosis of pneumonia (confirmed with a simple chest x-ray and/or thoracic computed tomography -CT-) by COVID-19 (confirmed with PCR in nasopharyngeal exudate and/or serological tests). Ultrasound was performed in the first 48 hours after hospital admission or transfer to our Service.

Following recommendations published jointly by the Spanish Society of Pneumology and Thoracic Surgery (SEPAR) and the Spanish Association of Respiratory Endoscopy (AEER)^22^, 6 areas were delimited in each hemithorax (Figure 1). In each area, all accessible intercostal spaces were explored, from medial to lateral and from inferior to superior. A score was assigned to each area according to the following scale (in the case of observing more than one in the same area, the highest score was assigned):

**Figure 1:**
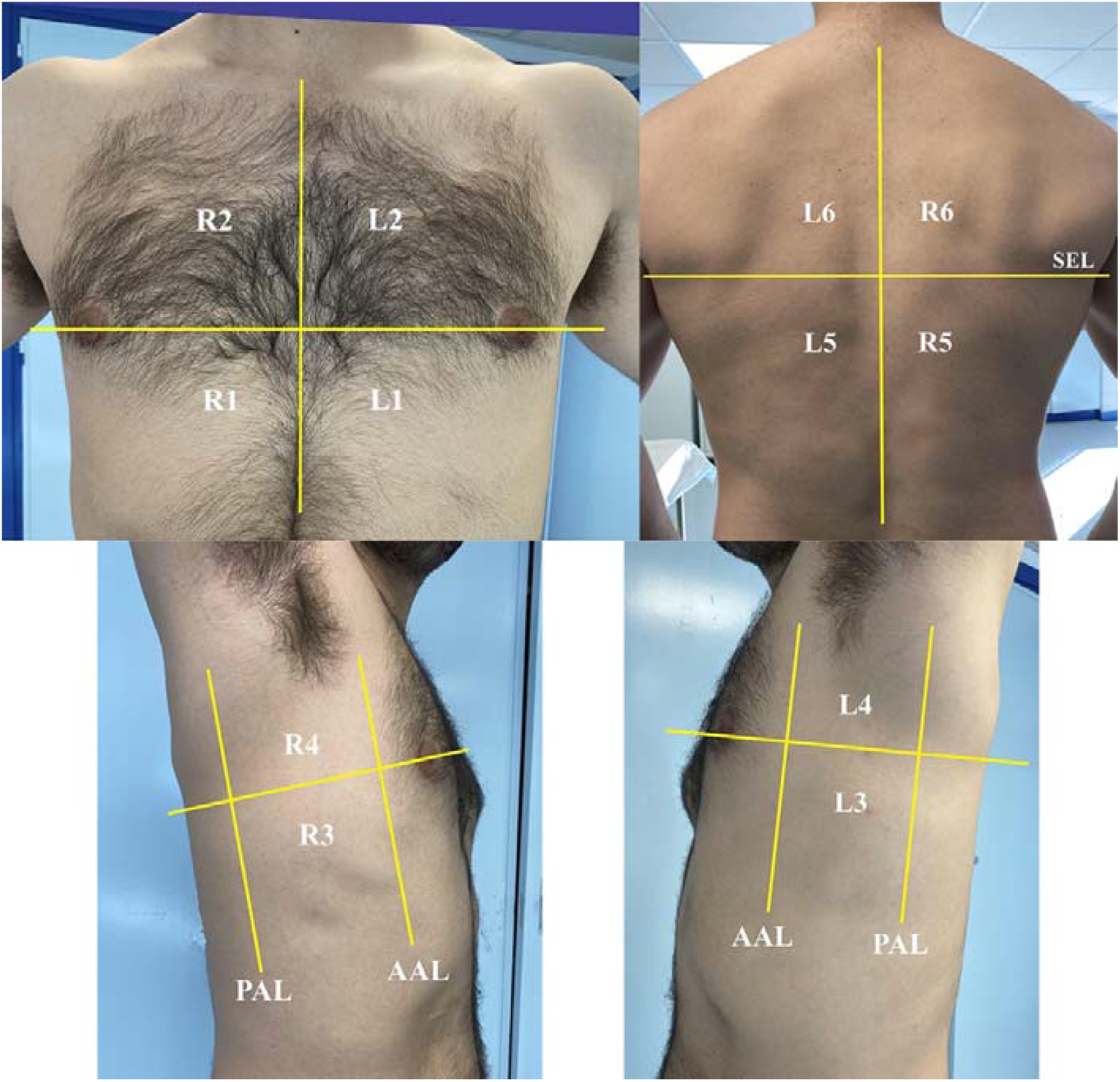
Areas for lung ultrasound SEL: subscapular line; AAL: anterior axillary line; PAL: posterior axillary line

- 0 points: A-pattern, defined by the existence of a regular and not thickened pleural line, with sliding sign, and the presence of A-lines.
- 1 point: B7-pattern, defined by the presence (in the same ultrasound field) of more than 3 B-lines with a separation of approximately 7 mm between them measured at the pleural line level.
- 2 points: B3-pattern, defined by the presence (in the same ultrasound field) of more than 3 B-lines with a separation of approximately 3 mm between them measured at the pleural line level, or the existence of confluent B-lines (white lung).
- 3 points: C-pattern, defined by the presence of an irregular or discontinuous pleural line, with small subpleural consolidations, or the existence of frank consolidation with air bronchogram and/or liquid bronchogram.

For purposes of analysis and comparisons, the following lung regions have been defined:

- Anterior region: R1 and R2 (right), L1 and L2 (left)
- Posterior region: R5 and R6 (right), L5 and L6 (left)
- Lateral region: R3 and R4 (right), L3 and L4 (left)
- Upper region: R2, R4 and R6 (right), L2, L4 and L6 (left)
- Lower region: R1, R3 and R5 (right), L1, L3 and L5 (left)

The score (lung score -LS-) was recorded for each area and for each region (sum of its areas), both unilaterally and bilaterally, as well as the total LS for each patient (minimum 0, maximum 36).

In 55 patients the ultrasound examination was performed while sitting, while in 8 was performed in the supine position; in the latter, the patient was temporarily placed in the right lateral position to explore L5 and L6 areas, and in the left lateral position to explore R5 and R6 areas.

The following data were also collected for each patient: age, sex, days elapsed since the onset of symptoms, PaO2/FiO2 ratio (PAFI) and need for non-invasive respiratory support (NIRS), which includes high-flow nasal cannula oxygen therapy, CPAP and noninvasive mechanical ventilation.

To perform the ultrasound, a C5-2 convex probe was used with the Lumify system using the lung configuration (Philips, Amsterdam, Netherlands) attached to a Galaxy Tab A 2019 tablet (Samsung Electronics, Seoul, South Korea). The ultrasound probe was protected with CIV-Flex cover (CIVCO Medical Instruments, Kalona, Iowa, USA) and the tablet with a clear plastic sleeve, which were discarded after each examination.

Statistical analysis has been performed with SPSS for Mac version 26 (IBM, New York, USA). Results of qualitative variables are expressed as percentages and absolute frequencies. For quantitative variables, mean and standard deviation are expressed. Comparison of discrete variables was carried out using the chi-square test, and that of independent means of quantitative variables using the Student’s t test. For comparison of multiple means, the block ANOVA with Tukey’s HSD post-hoc test was used. A p < 0.05 was considered statistically significant. For multivariate analysis, a binary logistic regression model with Hosmer-Lemeshow goodness of fit test was used. For the analysis of the ROC curves, sensitivity, specificity and Youden’s index were used.

## RESULTS

Patients’ demographic and clinical characteristics are presented in Table 1.

**Table 1:** **Demographic and clinical characteristics of patients**

|  | **Men** | **Women** | ***P*-value** |
| --- | --- | --- | --- |
| **No. of patients (%) (*n* = 63)** | 43 (68.3) | 20 (31.7) | 0.004 |
| **Age (y), mean ± SD** | 59.9 ± 14.0 | 64.0 ± 17.7 | 0.369 |
| **Days from symptoms onset, mean ± SD** | 11.3 ± 5.3 | 9.8 ± 5.8 | 0.332 |
| **PAFI, mean ± SD** | 300.8 ± 93.5 | 312.3 ± 93.9 | 0.655 |

PAFI: PaO<sub>2</sub>/FiO<sub>2</sub>; SD: standard deviation

### Ultrasound involvement

A-pattern was found in 251 out of 756 examined lung areas (33.2%), B7-pattern in 203 (26.8%), B3-pattern in 143 (19%) and C-pattern in 159 (21%).

#### Distribution of ultrasound findings

The distribution of ultrasound findings by pulmonary areas is shown in Table 2. We found ultrasound involvement (defined as the finding of a pattern other than A-pattern) in 8 ± 3.5 lung areas per patient (4.1 ± 1.7 in the right lung, 3.9 ± 1.9 in the left lung, *p* = 0.306). Involvement of at least one lung area was observed in all patients. In only 4 out of 63 patients (6.3%) the involvement was unilateral. Small unilateral pleural effusion was observed in 3 patients (4.8%).

**Table 2:** **Distribution of ultrasound findings by lung areas**

|  | **Right lung** |  |  |  |  |  |
| --- | --- | --- | --- | --- | --- | --- |
|  | **R1** | **R2** | **R3** | **R4** | **R5** | **R6** |
| **A-pattern** | 28 (44.4) | 30 (47.6) | 23 (36.5) | 26 (41.3) | 5 (7.9) | 10 (15.9) |
| **B7-pattern** | 19 (30.2) | 20 (31.7) | 13 (20.6) | 13 (20.6) | 9 (14.3) | 22 (34.9) |
| **B3-pattern** | 14 (22.2) | 10 (15.9) | 17 (27) | 16 (25.4) | 10 (15.9) | 15 (23.8) |
| **C-pattern** | 2 (3.2) | 3 (4.8) | 10 (15.9) | 8 (12.7) | 39 (61.9) | 16 (25.4) |
|  | **Left lung** |  |  |  |  |  |
|  | **L1** | **L2** | **L3** | **L4** | **L5** | **L6** |
| **A-pattern** | 31 (49.2) | 31 (49.2) | 23 (36.5) | 21 (33.3) | 8 (12.7) | 15 (23.8) |
| **B7-pattern** | 21 (33.3) | 21 (33.3) | 16 (25.4) | 18 (28.6) | 11 (17.5) | 20 (31.7) |
| **B3-pattern** | 10 (15.9) | 8 (12.7) | 13 (20.7) | 12 (19.1) | 7 (11.1) | 11 (17.5) |
| **C-pattern** | 1 (1.6) | 3 (4.8) | 11 (17.5) | 12 (19) | 37 (58.7) | 17 (27) |

Results are expressed as number of patients (%)

#### Anteroposterior distribution

the posterior region was affected in 118 out of 126 examined lungs (93.6%), the lateral one in 93 (73.8%) and the anterior one in 79 (62.7%). Only in 23 lungs (18.2%) a single region was affected: in 19 (15.1%) the posterior one, in 1 (0.8%) the lateral one, and in 3 (2.4%) the anterior one.

#### Craniocaudal distribution

the upper region was affected in 111 out of 126 examined lungs (88.1%) and the lower one in 120 (95.2%). Only in 13 lungs (10.3%) a single region was affected: in 11 (8.7%) the lower one and in 2 (1.6%) the upper one.

#### Degree of involvement

The mean total LS per patient was 15.3 ± 8.1 (range 1 to 33). The comparison of LS between lung regions is shown in Table 3.

**Table 3:** **Comparison of ultrasound involvement by lung regions**

| | Mean LS ± SD | *P*-value |
| --- | --- | --- |
| Right lung | 7.8 ± 4.1 | 0.377 |
| Left lung | 7.5 ± 4.5 | |
| Lower region | 8.4 ± 4.1 | < 0.001 |
| Upper region | 6.9 ± 4.5 | |
| Posterior region | 7.5 ± 3.1 | < 0.001* |
| Anterior region | 3.1 ± 2.8 | |
| Lateral region | 4.8 ± 3.5 | |

\*between any pair of data
LS: lung score; SD: standard deviation

### Correlation with gasometric severity

The mean of PAFI was 304.4 ± 93 (range 141 to 462). Table 4 shows the comparison of PAFI according to the presence or absence of ultrasound signs of consolidation (C-pattern) and the involvement or not of different lung regions.

**Table 4:** **Comparison of PAFI according to the presence of ultrasound signs of consolidation and the involvement of different lung regions**

|  | **Patients**<br>**n (%)** | **PAFI**<br>**Mean ± SD** | **P-value\*** |
| --- | --- | --- | --- |
| **Presence of consolidation:** |  |  |  |
| Yes | 52 (82.5) | 286.9 ± 90.9 | 0.001 |
| No | 11 (17.5) | 387.1 ± 49.4 | |
| **Involvement of anterior region:** |  |  |  |
| Yes | 45 (71.5) | 272.9 ± 87.3 | < 0.001 |
| No | 18 (28.5) | 383.3 ± 50.9 | |
| **Involvement of posterior region:** |  |  |  |
| Yes | 62 (98.4) | 302.5 ± 92.5 | 0.198 |
| No | 1 (1.6) | 424 |  |
| **Involvement of lateral region:** |  |  |  |
| Yes | 53 (84) | 289.5 ± 91.1 | 0.03 |
| No | 10 (16) | 383.8 ± 57.8 | |
| **Involvement of upper region:** |  |  |  |
| Yes | 59 (93.5) | 299.1 ± 93.0 | 0.04 |
| No | 4 (6.5) | 383.8 ± 53.1 | |
| **Involvement of lower region:** |  |  |  |
| Yes | 62 (98.4) | 302.5 ± 92.5 | 0.198 |
| No | 1 (1.6) | 424 |  |

\*for the comparison between means of PAFI

Correlation between PAFI and total LS shows r = -0.765 (p <0.001). Correlation between PAFI and LS of the different lung regions is shown in Figure 2, being especially striking with the LS of the anterior and superior regions.

**Figure 2:**
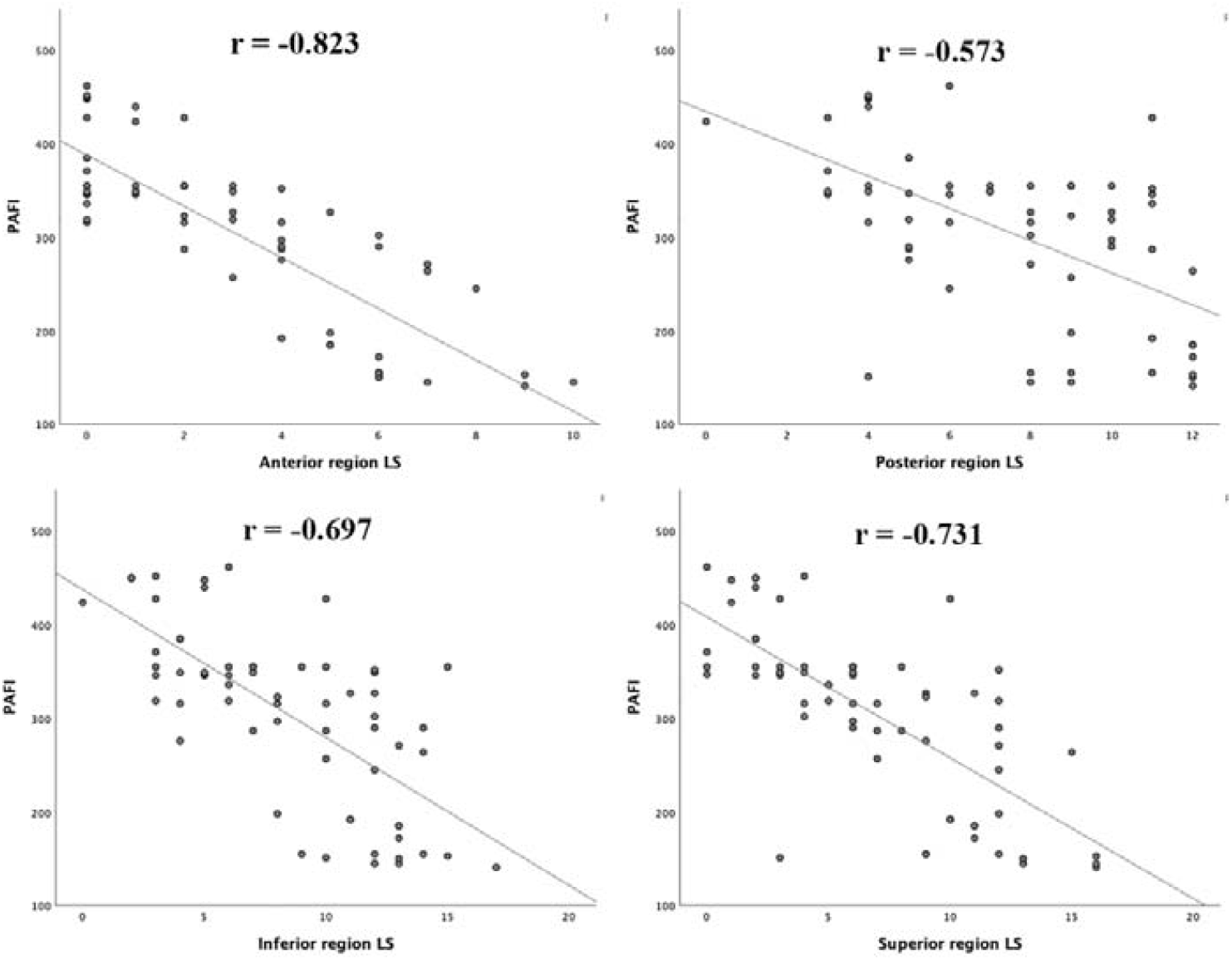
Correlation between PAFI and degree of ultrasound involvement of different lung regions LS: lung score; PAFI: PaO_2_/FiO_2_

### Prognostic value

Table 5 presents patients’ characteristics according to the need of NIRS, where we can see that, although patients who required NIRS had involvement of all lung regions, best discrimination occurs in the anterior one (*p* = 0.006).

**Table 5:** **Sociodemographic, clinical and lung ultrasound characteristics of patients according to the need for non-invasive respiratory support**

|  | **With NIRS** | **Without NIRS** | **P-value** |
| --- | --- | --- | --- |
| **No. of patients (%) (*n* = 63)** | 14 (22.2) | 49 (77.8) | - |
| **Man, N (%)** | 10 (71.4) | 33 (67.3) | 1.000 |
| **Age (y) (m ± SD)** | 73.9 ± 9.6 | 57.6 ± 14.6 | < 0.001 |
| **Days from symptoms onset (m ± SD)** | 14.0 ± 4.2 | 9.9 ± 5.5 | 0.012 |
| **PAFI (m ± SD)** | 196.5 ± 64.7 | 335.3 ± 75.3 | < 0.001 |
| **Superior region LS (m ± SD)** | 11.4 ± 3.7 | 5.6 ± 3.9 | < 0.001 |
| **Inferior region LS (m ± SD)** | 12.1 ± 2.3 | 7.4 ± 3.9 | < 0.001 |
| **Anterior region LS (m ± SD)** | 5.9 ± 2.5 | 2.2 ± 2.3 | < 0.001 |
| **Lateral region LS (m ± SD)** | 7.9 ± 2.1 | 3.8 ± 3.3 | < 0.001 |
| **Posterior region LS (m ± SD)** | 9.6 ± 2.3 | 6.9 ± 3.0 | 0.003 |
| **Right lung LS (m ± SD)** | 11.4 ± 2.7 | 6.8 ± 3.9 | < 0.001 |
| **Left lung LS (m ± SD)** | 12.1 ± 2.9 | 6.2 ± 4.0 | < 0.001 |
| **Total LS (m ± SD)** | 23.5 ± 5.3 | 13.0 ± 7.2 | < 0.001 |
| **Posterior region involvement, N (%)** | 14 (100) | 48 (98.0) | 1.000 |
| **Anterior region involvement, N (%)** | 14 (100) | 31 (63.3) | 0.006 |
| **Lateral region involvement, N (%)** | 14 (100) | 39 (61.9) | 0.065 |
| **Upper region involvement, N (%)** | 14 (100) | 45 (91.8) | 0.567 |
| **Lower region involvement, N (%)** | 14 (100) | 48 (98.0) | 1.000 |

LS: lung score; m: mean; N: number of patients; NIRS: non-invasive respiratory support; SD: standard deviation; PAFI: PaO<sub>2</sub>/FiO<sub>2</sub>

In a univariate logistic regression model for the risk of requiring NIRS, the significant variables were age (odds ratio -OR-: 1.096, 95% confidence interval -CI-: 1.037-1.158) and anterior region LS (OR: 1.809, CI: 1.303-2.510). In the multivariate model OR for age was 1.130, CI 1.041-1.277, while OR for anterior region LS was 2.159, CI 1.309-3.561.

Figure 3 shows ROC curves for the need of SRNI according to the LS of different lung regions. The ones with the best characteristics are total LS and anterior region LS. Table 6 shows sensitivity, specificity and Youden’s index of different cut-off points of total LS and anterior region LS for predicting the need of NIRS. The best cut-off points were total LS ≥ 19 and anterior region LS ≥ 4.

**Table 6:** **Sensibility, specificity and Youden’s index for the prediction of requiring NIRS at different cut-off points of lung score**

|  | **Sensibility (CI)** | **Specificity (CI)** | **YI** |
| --- | --- | --- | --- |
| **Anterior region LS $\geq 3$** | 13/14<br>0.929 (0.758 - 1) | 30/49<br>0.612 (0.466 - 0.759) | 0.541 |
| **Anterior region LS $\geq 4$** | 13/14<br>0.929 (0.758 - 1) | 35/49<br>0.714 (0.578 - 0.851) | 0.643 |
| **Anterior region LS $\geq 5$** | 10/14<br>0.714 (0.442 - 0.987) | 40/49<br>0.816 (0.698 - 0.935) | 0.530 |
| **Total LS $\geq 18$** | 12/14<br>0.857 (0.638 - 1) | 36/49<br>0.735 (0.601 - 0.867) | 0.592 |
| **Total LS $\geq 19$** | 12/14<br>0.857 (0.638 - 1) | 39/49<br>0.796 (0.673 - 0.919) | 0.653 |
| **Total LS $\geq 20$** | 12/14<br>0.857 (0.638 - 1) | 39/49<br>0.796 (0.673 - 0.919) | 0.653 |
| **Total LS $\geq 21$** | 11/14<br>0.786 (0.535 - 1) | 40/49<br>0.816 (0.698 - 0.935) | 0.602 |

CI: 95% confidence interval; LS: lung score; NIRS: non-invasive respiratory support; YI: Youden's index

**Figure 3:**
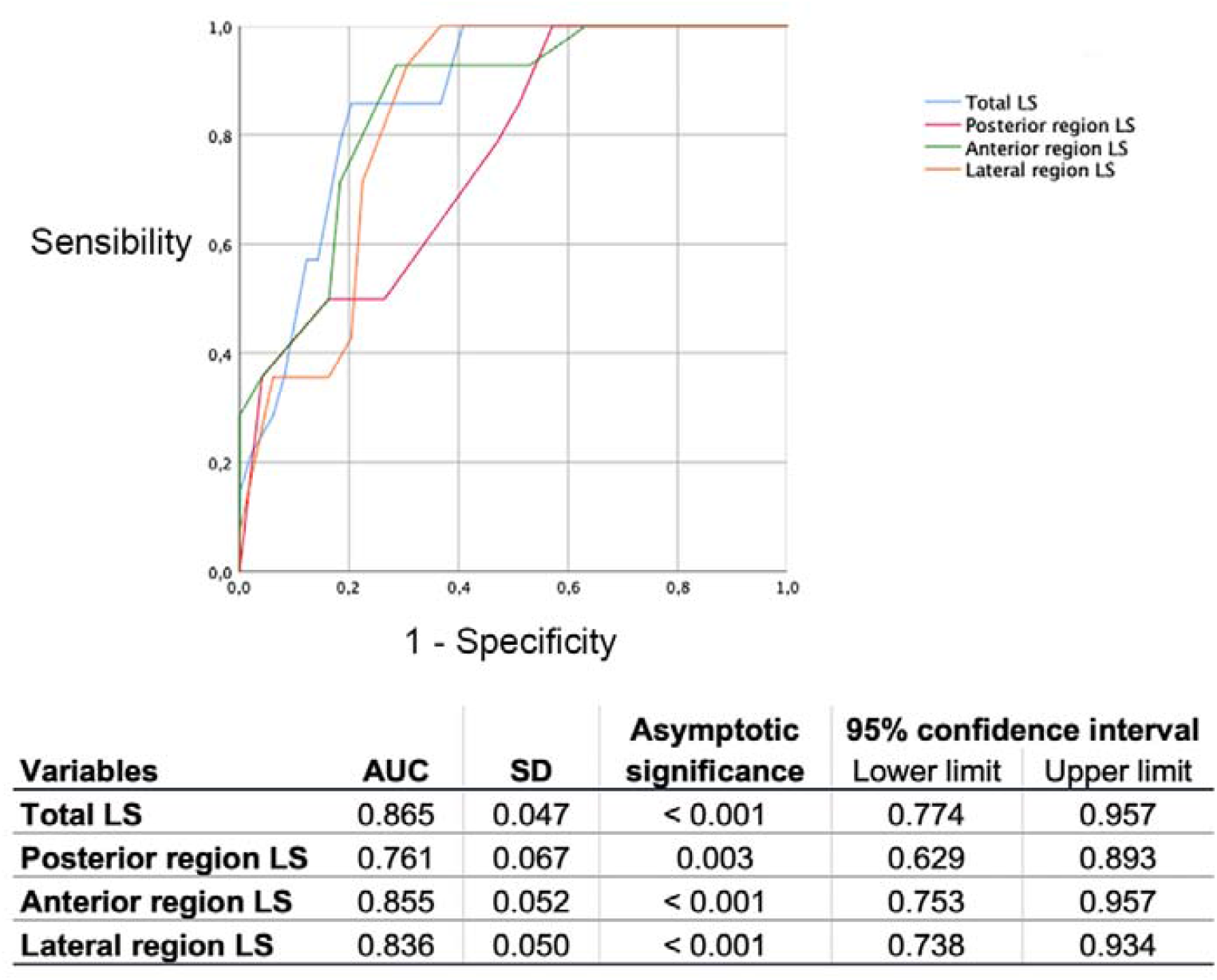
ROC curves for the need of NIRS according to the degree of ultrasound involvement of different lung regions NIRS: non-invasive respiratory support; LS: lung score; AUC: area under the curve, SD: standard deviation

## DISCUSSION

This study systematically describes the findings of pulmonary ultrasound in patients with COVID-19 pneumonia and identifies its prognostic value, since the LS of the anterior lung region correlates with gasometric severity and the need for NIRS. In our series, distribution by sex shows a significantly greater representation of men. This has not been observed in COVID-19 general population^5, 23^ although in some series that focus on patients with pneumonia, a higher proportion of men has been reported^24,25^, but not in others^26,27^.

### Ultrasound involvement

To calculate the LS of each area we have used the 4-point scale developed for the ultrasound evaluation of respiratory distress syndrome^28^, which is the one used by all authors to date, although recently Soldatti et al.^19^ made a slightly different standardization proposal specific for COVID-19 patients.

Most affected lung region is the posterior one, followed by the lateral one and the anterior one. Involvement is bilateral in most cases (93.7%) and finding of pleural effusion is rare (4.8%). These data are in line with that published in the series that analyse lung involvement both with chest CT^25-27,29^ and with ultrasound^11,15^.

There is no significant difference in ultrasound involvement between left and right lung. But there is, however, by lung regions, so that the lower region is affected more frequently than the upper one and its degree of involvement is also greater. The same occurs with the posterior region, which is affected more frequently than the anterior one and with a greater degree of involvement, which may perhaps help explain the improvement in oxygenation observed with pronation in many patients, both in classic distress^31^ and in COVID-19 pneumonia^31,32^.

### Gasometric severity

PAFI is one of the most objective and reliable parameters for assessing oxygenation status^33^, it is an important criterion for assessing the severity of pneumonia^34^ and defines severity in acute respiratory distress syndrome^35^. In our results, PAFI shows a good negative correlation with total LS, which is not surprising, but it is especially relevant that when examined by lung regions, the most significant negative correlation of PAFI was with LS of the anterior region. On the other hand, patients who have ultrasound involvement of the anterior region have a significantly lower mean PAFI than those who do not.

### Prognostic value

22.2% of patients required NIRS. When variables are entered into a multivariate logistic regression model to determine the risk of requiring NIRS, those that show statistical significance are age and LS of the anterior region.

Total LS and anterior region LS behave similarly in terms of characteristics of their ROC curves to predict the need for NIRS. When the possible cut-off points are analysed, total LS ≥ 19 and anterior region LS ≥ 4 show similar values. From a practical point of view, this may be important in some situations, because ultrasound examination of the anterior region requires less time and is more accessible than a complete ultrasound examination, especially in patients who cannot adopt the sitting position.

### Weakness and strength

The main weakness of this study is that not all patients admitted for COVID-19 pneumonia have been included, since the enormous caseload of these days prevented ultrasound examination for all of them. However, we believe that this fact does not cause an inclusion bias, since the decision to perform or not ultrasound depended only on the care burden of each day.

The main strength is that this is a prospective study and that the ultrasound examination was done with the operator unaware about patients’ radiological examinations and clinical situation, knowing only the diagnosis of COVID-19 pneumonia. This eliminates the part of subjectivity that may have the interpretation of ultrasound images.

## Conclusions

In conclusion, in COVID-19 pneumonia the ultrasound involvement is bilateral and heterogeneous. Pleural effusion is very rare. The degree of total ultrasound involvement shows a strong correlation with gasometric severity of pneumonia. The most affected lung regions are the posterior and the inferior ones. The anterior region, although usually more spared, is the one that implies a worse prognosis since its degree of involvement, along with age, predicts the need for non-invasive respiratory support. An anterior lung region lung score ≥ 4 has a high sensitivity and specificity to predict the need for non-invasive respiratory support.

## Data Availability

Data Availability: no

